# Modeling the USA Winter 2021 CoVID-19 Resurgence

**DOI:** 10.1101/2022.01.06.22268868

**Authors:** Genghmun Eng

## Abstract

The current USA 2021 CoVID-19 Winter Resurgence is modeled here with the same function used for analyzing prior USA CoVID-19 waves:

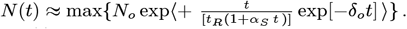

Here, *N*(*t*) gives the total number of CoVID-19 cases above the previous baseline, and *t*_*R*_ sets the initial *t*_*dbl*_ = *t*_*R*_ (ln 2) pandemic *t*_*dbl*_ doubling time. Larger *α*_*S*_ values indicate that uninfected people are improving their pandemic mitigation efforts, such as *Social Distancing* and *vaccinations* ; while *δ*_*0*_ *>* 0 accelerates the post-peak 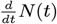 *tail-off*, and is empirically associated with *mask-wearing*. The pandemic wave end is when *N*(*t*) no longer increases.

The USA Summer 2021 resurgence results from our prior *medrxiv*.*org* preprints^*^ were used as a baseline. By 11/15/2021, an additional 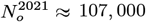 cases above baseline were found, signaling the USA Winter 2021 resurgence. This CoVID-19 wave is still in its *initial* stages. Presently, our analysis indicates that this CoVID-19 wave can infect virtually all susceptible persons; just like the *initial* stage of the USA Summer 2021 resurgence. Data up through 12/30/2021 gives these paremeter values:

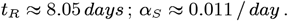

These values are identical to the prior 2020 USA Winter Resurgence results. Also, the 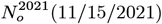 and the 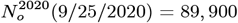 values are similar. However, while the Winter 2020 Resurgence showed a significant *mask-wearing* effect 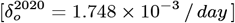, this *initial* USA Winter 2021 Resurgence shows practically no *mask-wearing* effects 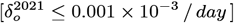. If *mask-wearing* were to quickly rise to the Winter 2020 levels, it would give these projected totals:

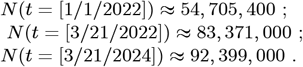

More robust *mask-wearing* and enhanced *Social Distancing* measures could further reduce these values (*with 3 Figures*).

## 1 Introduction

Each new USA CoVID-19 wave usually starts with a sharp rise in the total number *N*(*t*) of new cases. The time evolution for each of these waves has been successfully modeled^**1–6**^ using this basic function for *N*(*t*):

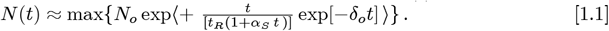

Equation [1.1] represents the USA Winter 2021 CoVID-19 Resurgence cases above a baseline that is set by the USA Summer 2021 Resurgence from our prior *medrxiv*.*org* preprints^**5–6**^. The standard **SEIR** (**S**usceptible, **E**xposed, **I**nfected, and **R**ecovered or Removed) epidemiology models all start with an exponential growth (*R*_*o*_ *>* 1) or decay (*R*_*o*_ *<* 1):

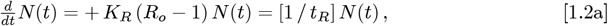

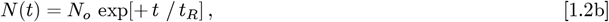

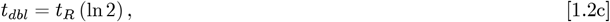

where *t*_*R*_ *>* 0 sets the initial growth rate, and *t*_*dbl*_ sets the initial *N*(*t*) doubling-time. Virtually all **SEIR** models are *local* models, with *R*_*o*_ representing the average number of people who will become infected by an ailing person during the course of their illness.

Given a total population of *N*_*ALL*_, the *uninfected population U*(*t*) is:

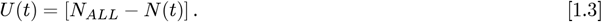

Using Eq. [1.1] implicitly assumes that *N*(*t*) *<< N*_*ALL*_ for all times of interest, so that pandemic saturation effects can be ignored. In general, **SEIR** models do not explicitly consider what the *U*(*t*) *uninfected population* may be doing in response to the pandemic, prior to becoming infected. In contrast, Eq. [1.1] was developed as a *non-local* extension of **SEIR** models, to explicitly take into account what the *uninfected population*, as a whole, is doing to mitigate pandemic spread.

Since Eq. [1.1] is empirically based, it does not predict when each new CoVID-19 wave will start, or what biological and social circumstances are causing the new wave. As a result, the *t* = 0 point for each wave is usually set by when the resurgence is first easily identified, with *N*(*t* = 0) = *N*_*o*_ being the number of cases above baseline at that time. But once the CoVID-19 wave becomes established, Eq. [1.1] appears to successfully predict its time evolution.

As given in our prior preprints^**1–6**^, the parameter *α*_*S*_ measures the combined effect of virtually all large-scale pandemic mitigation efforts. These include *Social Distancing* requirements, such as minimum separation distances and decreased allowable occupancy; along with lockdowns, “stay at home” orders, school closures, and restrictions on business operations; as well as the impact of large-scale vaccination efforts. The *δ*_*o*_ *>* 0 parameter accelerates the post-peak 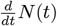 *tail-off*, and our prior work^**3–6**^ shows that it is empirically associated with *mask-wearing*.

The calculated end for each pandemic wave occurs when *N*(*t*) in Eq. [1.1] for the total number of cases first stops increasing, which makes the final stages of calculated post-peak 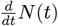 *tail-off* inaccurate. However, the pandemic wave is substantially over by then, assuming that no follow-on resurgence occurs.

## 2 The USA Winter 2021 Resurgence

Our CoVID-19 modeling using the same few parameters has been successful at predicting the time evolution of each prior USA CoVID-19 wave^**1–6**^. This result shows that the **response** of the *U*(*t*) *uninfected population* was similar for each wave, even if different dominating factors drove each new resurgence.

Deviations of ∼107, 000 extra cases above the USA Summer 2021 baseline were observed by 11/15/2021. Thus, the USA is now in the *initial* stages of a Winter 2021 resurgence, as shown in *Figure 1*. Our calculations indicate that this CoVID-19 wave presently can infect virtually all susceptible people; a result that is similar to the *initial* stage of the USA Summer 2021 wave^**5**^.

**Figure 1.**
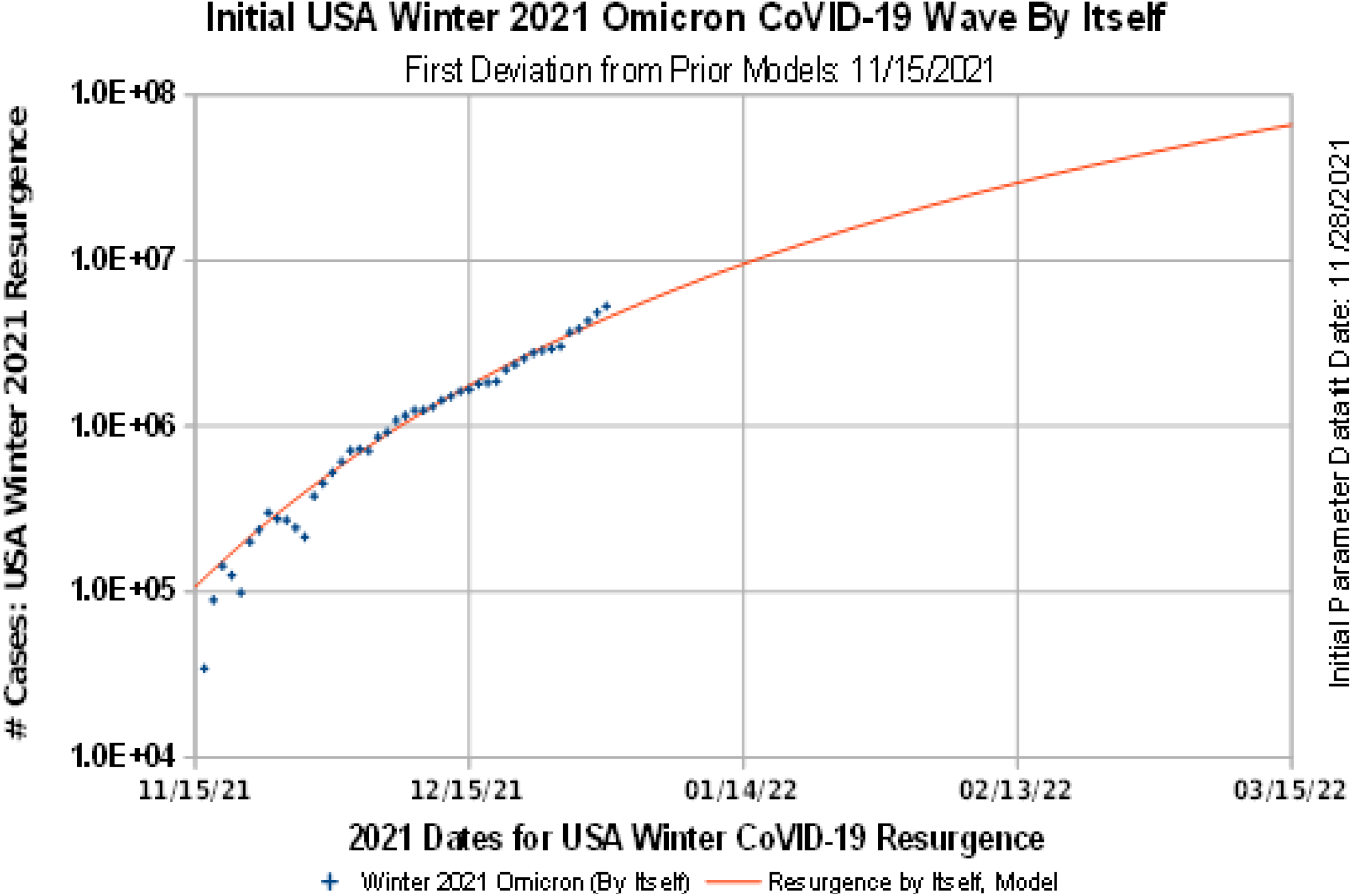
The USA CoVID-19 Winter 2021 Resurgence, By Itself.

In addition, two of the three Eq. [1.1] parameter values, as determined using the data up through 12/30/2021, were found to be identical to the 2020 USA Winter Resurgence values as follows:

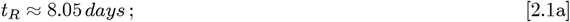

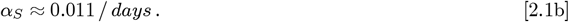

Both CoVID-19 Winter Resurgences also have similar *N*_*o*_ values:

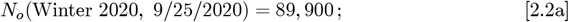

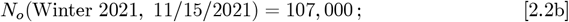

indicating that both waves started similarly. However, the Winter 2020 Resurgence was associated with a non-negligible amount of *mask-wearing*, as measured by *δ*_*o*_ in Eq. [1.1]. In contrast, this *initial* stage of the USA Winter 2021 wave is associated with having virtually no *mask-wearing* effects:

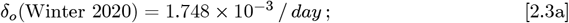

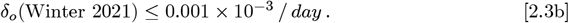

In *Figure 2*, the {*N*_*o*_; *t*_*R*_; *α*_*S*_; *δ*_*o*_} parameter values for this Winter 2021 Resurgence are compared to each of the prior USA CoVID-19 waves. If *mask-wearing* were now to quickly rise to the Winter 2020 levels, without changing the other Winter 2021 Resurgence parameters, the total number of projected USA CoVID-19 cases would be:

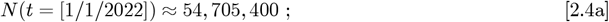

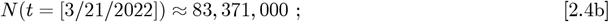

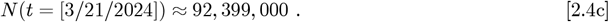

**Figure 2.**
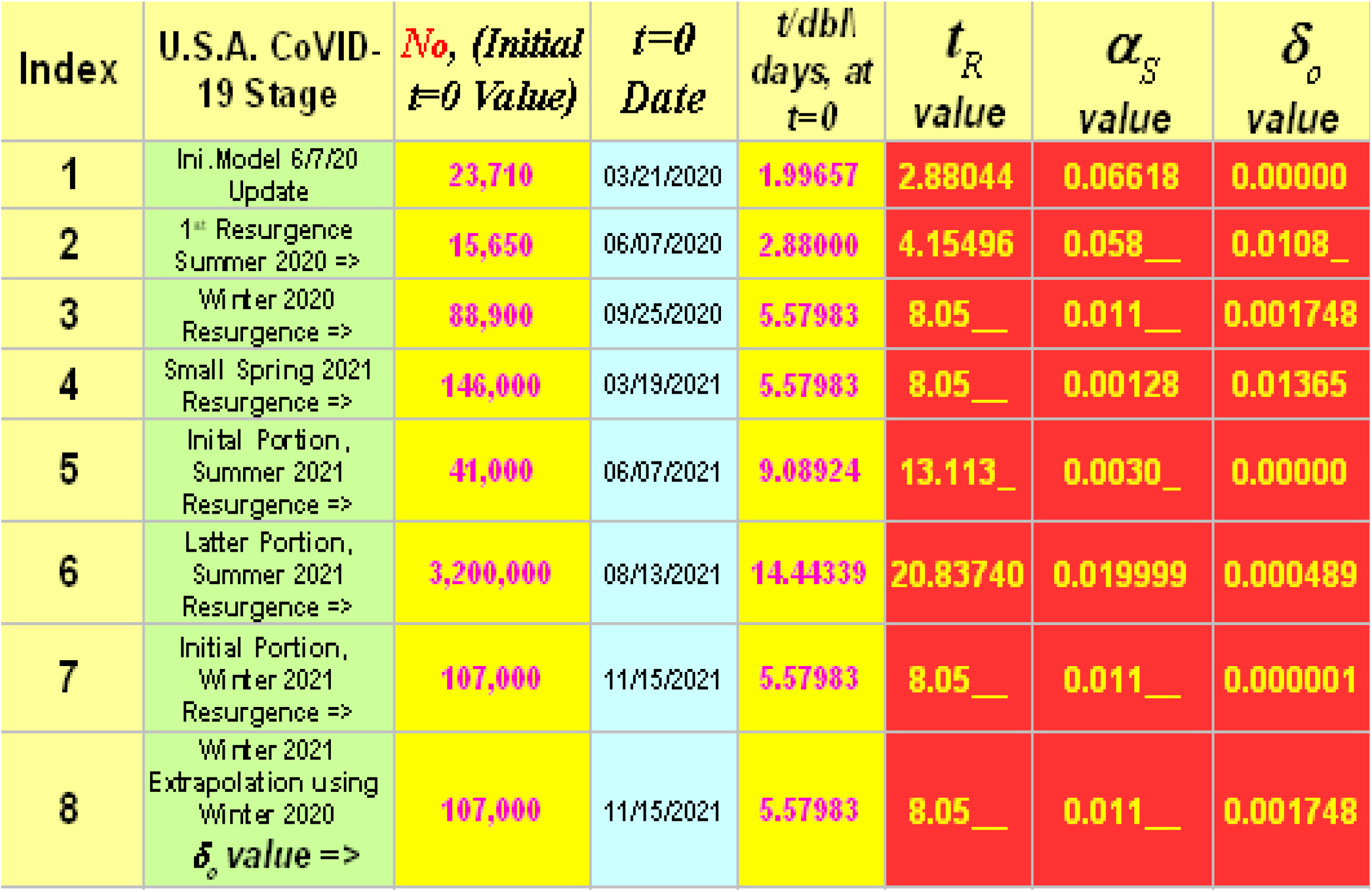
Summary of CoVID-19 Model and Parameter Values.

Without *mask-wearing* [*δ*_*o*_ *≈* 0], the 3*/*21*/*2022 projection would change to:

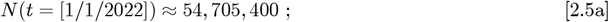

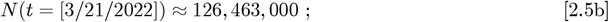

with the calculated *N*(*t*) for [3*/*21*/*2024] substantially exceeding the *N*_*ALL*_ total US population. At that point the Eq. [1.1] assumption that *N*_*ALL*_ is large enough so that *N*(*t*) *<< N*_*ALL*_ for all times of interest, would no longer be valid. The USA would be in a pandemic saturation stage, where practically everybody is or was infected.

*Figure 3* shows our model predictions vs CoVID-19 data for the entire pandemic, from March 2020 through January 2022. Whenever the pandemic appeared to be beaten down, restrictions were relaxed, CoVID-19 cases increased, new CoVID-19 variants appeared, and the pandemic rose up again, almost with every season.

**Figure 3.**
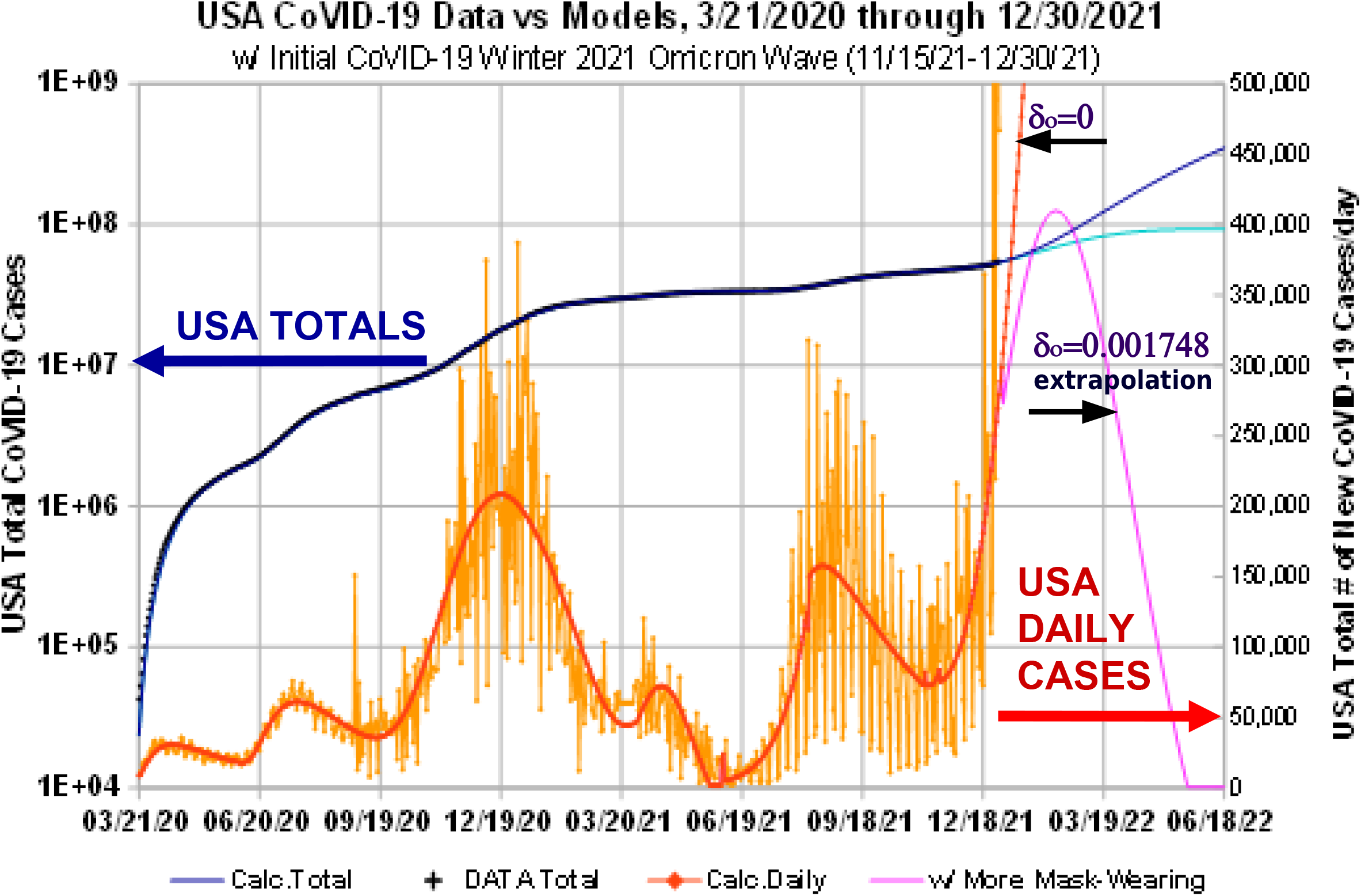
USA CoVID-19 Totals: 3/21/2020 through 12/30/2021.

The daily number of new cases *dN*(*t*) */ dt* in *Figure 3* has peaks for the initial Spring 2020 pandemic; a Summer 2020 resurgence; the long Winter 2020 Resurgence; a small uptick in Spring 2021; the USA Summer 2021 Resurgence; and now the USA Winter 2021 Resurgence.

In addition, *Figure 3* shows Winter 2021 Resurgence projections for two different *δ*_*o*_-values. One projection is the best datafit to date (12/30/2021), and uses *δ*_*o*_ ≤ 0.001 × 10^−3^ */ day*. The other projection is how the Winter 2021 Resurgence would progress, if *δ*_*o*_ were to suddenly increase to the prior Winter 2020 Resurgence value of *δ*_*o*_ = 1.748 × 10^−3^ */ day*.

## 3 Summary

It is January 2022, and the USA is in the midst of the *initial stages* of a CoVID-19 Winter 2021 Resurgence. Using the USA CoVID-19 Summer 2021 Resurgence as a baseline^**5–6**^, the latest CoVID-19 wave was modeled using:

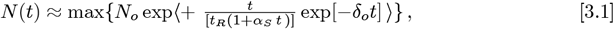

with those results shown in *Figure 1*. This function was used to analyze each previous USA CoVID-19 wave^**1–6**^. Since the same few parameters successfully apply to all these USA CoVID-19 waves, with only different *t* = 0 starting points and {*N*_*o*_; *t*_*R*_; *α*_*S*_; *δ*_*o*_} parameter values allowed, it shows that the **response** of the *U*(*t*) *uninfected population* has been similar for each CoVID-19 wave, even if different factors were driving each new resurgence.

This effect is best seen by comparing the parameters derived for this *initial stage* of the USA CoVID-19 Winter 2021 Resurgence to the prior USA CoVID-19 Winter 2020 Resurgence, as shown in *Figure 2*. Both have similar {*N*_*o*_; *t*_*R*_; *α*_*S*_} parameter values, indicating the USA population is in a similar position with respect to CoVID-19 in both cases. No doubt, the large number of Winter 2021 CoVID-19 Resurgence cases is being driven by infections from the most recent *Omicron* CoVID-19 variant.

However, back on Jan. 7, 2021, during the Winter 2020 Resurgence, only about ∼ 3% of the USA population had received any vaccinations, with only ∼0.3% being fully vaccinated. At that time, the *U*(*t*) *uninfected population* was substantially unvaccinated. In contrast, by Jan. 2022, a substantial fraction of the *U*(*t*) *uninfected population* was considered fully vaccinated.

Given the known capacity for the *Omicron* CoVID-19 variant to infect *fully vaccinated* persons, the similarity in the {*N*_*o*_; *t*_*R*_; *α*_*S*_} parameters between these two USA Winter Resurgences, shows that the USA *vaccinated population* is in a similar position to the USA *unvaccinated population* a year ago. Such a result brings CoVID-19 infection closer to becoming *endemic*. The ‘silver lining’ to this dour cloud is that those who are vaccinated appear to have much less severe CoVID-19 infection impacts.

One significant difference between this *initial stage* of the USA CoVID-19 Winter 2021 Resurgence and the prior USA CoVID-19 Winter 2020 Resurgence is that the *δ*_*o*_-parameter associated with *mask-wearing* was significantly higher in 2020, compared to this *initial stage* of the USA CoVID-19 Winter 2021 Resurgence, which showed *δ*_*o*_ ≈ 0 result.

If *δ*_*o*_ *≈* 0 persists, along with the present {*N*_*o*_; *t*_*R*_; *α*_*S*_} values, this CoVID-19 Winter 2021 Resurgence will have the capacity to infect nearly everyone who is not practicing the strictest CoVID-19 prevention protocols, with the resulting *N*(*t*) projections for *δ*_*o*_ ≈ 0 given in the prior section. Significantly increased *mask-wearing* and enhanced *Social Distancing* measures are needed to prevent these high levels of USA CoVID-19 projected cases from actually occurring.

## Data Availability

All data produced in the present work are contained in the manuscript.

## Footnotes

* *(10*.*1101_2021*.*08*.*16*.*21262150; 10*.*1101_2021*.*10*.*15*.*21265078)*

## References

1. https://medrxiv.org/cgi/content/short/2020.05.04.20091207v1 “Initial Model for the Impact of Social Distancing on CoVID-19 Spread”

2. https://medrxiv.org/cgi/content/short/2020.06.30.20143149v1 “Orthogonal Functions for Evaluating Social Distancing Impact on CoVID-19 Spread”

3. https://medrxiv.org/cgi/content/short/2020.08.07.20169904 “Model to Describe Fast Shutoff of CoVID-19 Pandemic Spread”

4. https://medrxiv.org/cgi/content/short/2020.09.16.20196063 “Initial Model for USA CoVID-19 Resurgence”

5. https://medrxiv.org/cgi/content/short/2021.08.16.21262150 “The IHME vs Me: Modeling USA CoVID-19 Spread, Early Data to the Fifth Wave”

6. https://medrxiv.org/cgi/content/short/2021.10.15.21265078 “Updated Model for the USA Summer 2021 CoVID-19 Resurgence”

